# Algorithm Refinement in the Non-Invasive Detection of Blood Glucose via Bio-RFID™ Technology^1^

**DOI:** 10.1101/2023.05.25.23290539

**Authors:** Dominic Klyve, Kaptain Currie, James H. Anderson, Carl Ward, David Schwarz, Barry Shelton

## Abstract

Diabetes Mellitus (DM) is a highly prevalent and increasingly common disorder that can have dire health consequences if not properly managed. Managing DM involves monitoring blood glucose levels, which can be cumbersome and invasive, and adherence to this practice is poor. We present a validation for a novel sensor designed to measure blood glucose (BG) non-invasively using Radio Frequency (RF) waves. In this n=5 study, we trained a Light Gradient-Boosting Machine (lightGBM) model to predict BG values using 1,555 observations from over 130 hours of data collection from five participants, where an observation is defined as data collected from 13 Bio-RFID sensor sweeps paired with a single Dexcom G6® value. Using this model, we predicted BG in the test set with a Mean Absolute Relative Difference (MARD) of 12.7% in the normoglycemic range and 14.0% in the hyperglycemic range. Overall, 70.7% of the estimates fell within 15% of the reference value, and 79.1% fell within 20% of the reference value. While this is a small participant sample, these strong initial results indicate the efficacy of this technique, and that with further refinement and more data, there is promise to achieve a clinically relevant level of accuracy.

## 1. Introduction

The prevalence of Diabetes Mellitus (DM) is staggering, affecting over 500 million individuals globally. DM is a condition characterized by high blood glucose (BG) that can result in severe long-term health consequences. To mitigate the risks associated with DM, it is crucial to regulate BG levels through lifestyle modifications, oral medications, and/or insulin injections^1^. To monitor BG, patients often rely on invasive portable measurement devices that require frequent finger pricking and disposable test strips, which are painful, and generate both ongoing expenses and biomedical waste. In some cases, patients may need up to ten BG measurements per day, which can be exceedingly uncomfortable and interfere with manual tasks such as typing, craftsmanship, or artistry.

Indeed, adherence with the practice of daily monitoring of BG is poor. A study of data from the National Health and Nutrition Examination Survey (NHANES) found that 29% of patients on insulin, 65% of patients on oral agents, and 80% of patients using diet alone had never monitored their blood glucose. Moreover, only 39% of patients using insulin and only 5-6% of those using oral agents had measured their BG daily^2^. Adherence to monitoring is fundamental to effective treatment. Additionally, contaminated consumables such as test strips and needles must be disposed of correctly to avoid potential health hazards related to the transmission of blood-borne diseases. Although modern continuous and less invasive glucose monitors (CGMs) exist, these devices are not without limitations and come with the additional cost of regular replacement and the discomfort of probe insertion. Hence, the development of a portable, non-invasive, and reliable point-of-care device for measuring BG is imperative.

One technique for non-invasive monitoring of glucose that has shown promise in other studies involves Radio Frequency (RF)/microwave detection^3^. This technique relies on the fact that BG affects the dielectric properties of the blood, which in turn changes the way that microwaves behave as they pass through it^4^. Most studies using such techniques have used devices that operate at a specific frequency in the 1 – 6 GHz range^5–10^, though some researchers have reported using a range of different frequencies^11^. Results from those studies have demonstrated that there is promise in this approach, though clinical precision has been illusive^12^.

In this paper, we describe our efforts to predict BG using data from five participants collected with a new type of sensing device that rapidly scans through a wide band of RF frequencies and records values detected at each frequency over a period of time. Note that in this context, “predict” is used in the machine learning sense – a “prediction” is the machine learning model’s estimate of a participant’s BG – and does not refer to predicting *future* values of BG. We use readings of a Dexcom G6®, a popular continuous glucose monitor (CGM), as a proxy for BG. Using a Light Gradient-Boosting Machine (lightGBM) model, we predict values of the Dexcom G6. As is typical in glucose measurement studies, accuracy of predictions was measured using the Mean Absolute Relative Difference (MARD). We demonstrate an overall MARD of 12.9%, which is within the range of FDA-approved CGM devices.

## 2. Methods

### 2.1 Overview of Participants

The study was approved by Core Human Factors IRB [IORG0007854; IRB Registration # IRB00009432], and all participants provided verbal and written informed consent to participate in the study. Five healthy adults (two female, three male) aged 29 – 61 participated in the study. Participants had no clinical history or diagnosis of diabetes or other significant medical conditions that could interfere with data collection or BG more broadly.

### 2.2 Data collection

Data were collected from each participant once each day in a research lab located in Seattle, Washington, up to a total of 12 days of glucose data collection. The research lab used for this study contains two testing rooms, each with a testing armchair equipped with a patented Know Labs Bio-RFID sensor built into each arm of the chair, allowing participants to simply rest their arms on the chair for the duration of the test.

The sensor consists of a Printed Circuit Board Assembly (PCBA) that generates RF signals and measures received power after passing those signals through an Antenna Array. Figure 1 shows an engineering diagram of the sensor and antenna array. There is a transmit (Tx) amplifier to boost the signal, and then the RF signal is routed through a switch matrix that allows for it to be sent to any one of the four supported antenna elements, or optionally through an onboard fixed-attenuation path called the Calibration Path that allows the system to test itself and provide a known benchmark.

**Figure 1:**
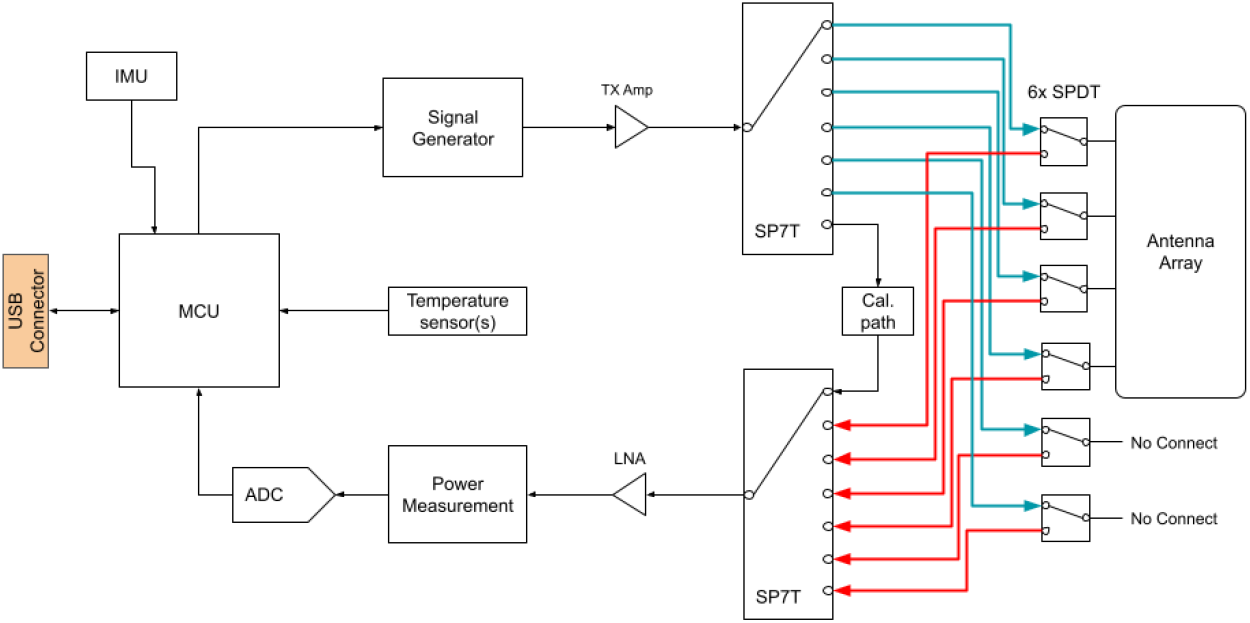
System architecture diagram of Know Labs Bio-RFID™ sensor

Roughly 24 hours before the start of testing, participants were fitted with a Dexcom G6 on the posterior of the upper left arm. Participants were asked to fast for at least 90 minutes before testing. During data collection, participants sat in the testing chair with the left forearm placed on the antenna of the sensor. Participants made efforts to minimize body movement for the duration of the test. Participants were seated 10 minutes prior to the start of the test. Glucose values were recorded from the Dexcom G6 every 5 minutes (i.e. the sampling rate of the reference device) for the duration of the test. The Dexcom G6 values collected during the first 30 minutes of the test were considered to be the individual’s “baseline” BG. At 30 minutes, the participant consumed 37.5 g of liquid D-Glucose (Azer Scientific Glucose Drink #10-LL-075). Testing continued until the participant’s BG returned to baseline for 30 minutes or until a maximum test time of 3.5 hours had passed.

Data were collected continuously from the Bio-RFID sensor using sweeps across the 500 MHz – 1500 MHz range at 0.1 MHz intervals, so each sweep collected data on 10,001 frequencies. Each sweep took approximately 22 seconds, including a one second pause between sweeps.

### 2.3 Data preprocessing

The dataset used in this analysis contained 1,555 Dexcom G6 values collected at 5-minute intervals, and 22,615 Bio-RFID sensor sweeps collected at 22-second intervals. In order to minimize noise in the data and to reduce the number of variables passed to the machine learning model, the Bio-RFID data were grouped in two ways. We downsampled the temporal domain by taking the mean of the five minutes of data (consisting of 13 frequency sweeps) leading up to a Dexcom G6 measurement. We downsampled the frequency domain by taking the mean of each set of 250 consecutive frequencies, so that the model received data in 25MHz intervals rather than the 0.1 MHz intervals in the original data.

After preprocessing, the final dataset used in model development contained 1,555 observations, each comprising data from 13 Bio-RFID sensor sweeps paired with a single Dexcom G6 value. According to the reference device, participants’ BG during the tests ranged from 65 – 278 mg/dL (3.61 mmol/L – 15.4 mmol/L), with 88.1% of values in the normoglycemic range (defined as 70 – 180 mg/dL or 3.89 mmol/L – 10 mmol/L), 11.6% in the hyperglycemic range (over 180 mg/dL or 10 mmol/L), and 0.3% in the hypoglycemic range (under 70 mg/dL or 3.89 mmol/L).

### 2.4 Model architecture and training

The primary goal of this work was to develop a machine learning model that can predict values of the Dexcom G6 using the Bio-RFID data. We employed an 80-20 training-test split; a validation set was not used due to the relatively small size of the dataset, in order to maximize the number of observations the model had to train on, and thus all hyperparameter tuning was done entirely on the training set.

Given that this dataset is relatively small, factors that could contribute to variance were stratified to avoid creating a test dataset that was not representative of the training set. The training and test datasets were stratified by participant (n=5), Bio-RFID sensor (2 devices), and glycemic status using scikit-learn’s “train_test_split” function. That is, the 80% of data were randomly selected for the training dataset, with the constraint that it contained equal representation from each participant, device, and glycemic status.

We elected to train a model based on random forests. In particular, we employed a Light Gradient-Boosting Machine (lightGBM) model, implemented in the lightGBM package^13^ (version 3.3.5) in Python (version 3.10.11). LightGBM models were developed by Ke et al. in 2017, and make very few assumptions about the structure of the data, allowing significant flexibility in creation of the final model.

The lightGBM model was trained on the training dataset with MARD as the loss function. *L1, L2*, and feature fraction penalties were applied to avoid overfitting. Hyperparameter tuning was conducted on the penalty terms, and the lowest MARD achieved was taken for the final model, resulting at *L1* = .4, *L2* = .4, and feature fraction = .5. A sample tree taken from the resulting lightGBM model is shown in Figure 2.

**Figure 2:**
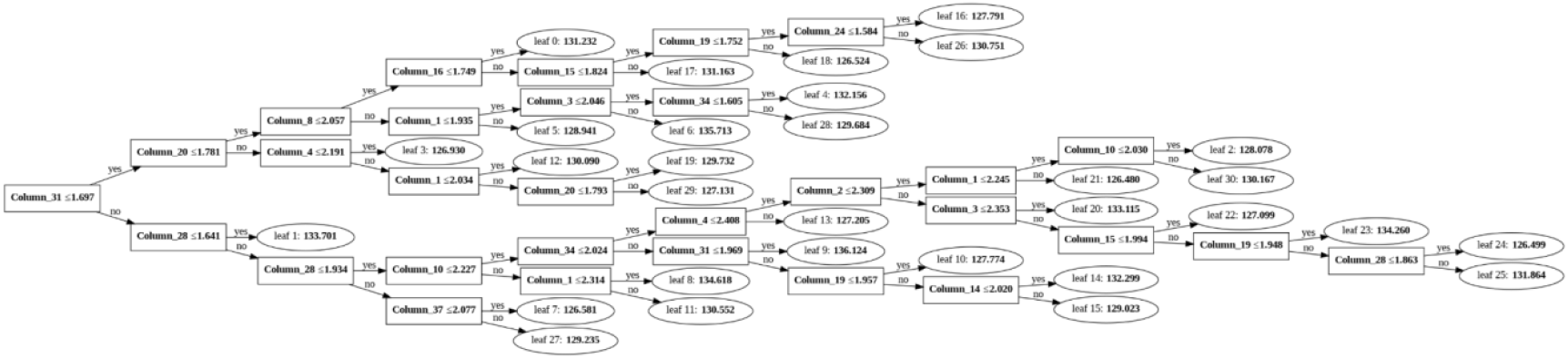
Visualization of a sample tree from the lightGBM architecture

### 2.5 Performance Metrics

We evaluated the model’s performance using the Mean Absolute Relative Difference (MARD). The Absolute Relative Difference (ARD) is the absolute value of the difference between the Dexcom G6 value and the value predicted by the model, divided by the reference (Dexcom G6) value. Thus, for example, if the model predicted a value of 86 while the Dexcom value was 90, the ARD would be (90-86)/90 ≅ 4.4%. The MARD is simply the mean of these ARDs across the entire dataset in question.

Other metrics considered for performance evaluation included the Mean Absolute Error (MAE), the proportion of predicted values that fell within 15% of the reference values for blood glucose (±15%), the proportion of predicted values that fell within 20% of the reference values for blood glucose (±20%), a Clarke Error Grid analysis, and a Surveillance Error Grid^14,15^. The “percent within threshold” metrics are given by the FDA, which requires approved glucose monitors to be accurate within 15% of the reference value 95% of the time, and within 20% of the reference value 99% of the time^16^. To contextualize these accuracy metrics in a way that is specific to this unique dataset, we calculated empirical chance for each metric. Empirical chance was calculated by randomly shuffling the test dataset’s reference values and comparing the model’s predictions for each observation compared against these values. This approach allows us to keep the true distribution of our test dataset’s reference values when calculating chance. Results were also broken down by each factor used for stratifying the test set, i.e. glycemic level, Bio-RFID sensor, and participant.

## 3. Results

### 3.1 LightGBM model compared against empirical chance

After the model was trained, inference was conducted on the test dataset, which contained 311 observations. Each blood glucose level predicted by Bio-RFID was then compared against the Dexcom G6 reference device’s BG measurement, with results shown in Table 1. In each metric, our model performed significantly better than the empirical chance model. The MARD in the test dataset was significantly lower than empirical chance (*t*=11.978, *p*<.001). The MAE in the test dataset was also significantly lower than empirical chance (*t*=10.860, *p*<.001). Via a proportion test, there were significantly more values that fell within 15% of the target value (*z*=8.995, *p*<.001) and significantly more values that fell within 20% of the target value (*z*=8.533, *p*<.001) than empirical chance.

**Table 1:** Accuracy of the predictions in the test dataset for both our model and empirical chance. Error values on the MARD and MAE give the 95% t-Confidence interval. Error bars on the ±15% and ±%20 give the 95% z-Confidence interval for proportions.

|  | MARD (%) | MAE (mg/dl) | ±15% | ±20% |
| --- | --- | --- | --- | --- |
| **LightGBM model** | 12.9 ± 1.4 | 16.2 ± 1.7 | 70.7% ± 5.1% | 79.1% ± 4.5% |
| **Empirical Chance** | 29.8 ± 1.3 | 37.6 ± 1.6 | 29.9% ± 5.1% | 40.5% ± 5.5% |
| **P value for difference** | $p<0.001^{***}$ | $p<0.001^{***}$ | $p<0.001^{***}$ | $p<0.001^{***}$ |

### 3.2 Comparing the training and test datasets

Despite precautions taken for overfitting described in Section 1.3, there is some evidence that overfitting occurred in the training dataset. In particular, the results were better in the training dataset than in the test dataset, as seen in Table 2.

**Table 2:** Results compared between training and test datasets. Error bars on the MARD and MAE give the 95% *t*-Confidence interval. Error bars on the ±15% and ±20% give the 95% *z*-Confidence interval for proportions.

|  | Samples | MARD (%) | MAE (mg/dl) | ±15% | ±20% |
| --- | --- | --- | --- | --- | --- |
| **Training** | 1244 | 6.0 ± 0.3 | 7.8 ± 0.5 | 92.7% ± 1.5% | 97.0% ± 1.0% |
| **Test** | 311 | 12.9 ± 1.4 | 16.2 ± 1.7 | 70.7% ± 5.1% | 79.1% ± 4.5% |

### 3.3 Comparing results across difference glycemic ranges

There were insufficient observations in the hypoglycemic range in the test dataset, and thus only normoglycemic and hyperglycemic ranges were compared. For a detailed comparison, see Table 3. The MAE in the normoglycemic range was significantly better than empirical chance (*t*=10.354, *p*<.001). In the hyperglycemic range, there was a tendency (*t*=1.822, *p*<.078) to a better MAE than empirical chance.

**Table 3:** Results broken down by glycemic status. Error bars on the MARD and MAE give the 95% *t*-Confidence interval. Error bars on the ±15% and ±20% give the 95% *z*-Confidence interval for proportions.

|  | Samples | MARD (%) | MAE (mg/dl) | ±15% | ±20% |
| --- | --- | --- | --- | --- | --- |
| **All observations** | 311 | 12.9 ± 1.4 | 16.2 ± 1.7 | 70.7% ± 5.1% | 79.1% ± 4.5% |
| **Hypoglycemic (<70 mg/dl)** | 1 (0.3%) | n/a | n/a | n/a | n/a |
| **Normoglycemic (70 - 180 mg/dl)** | 274 (88.1%) | 12.7 ± 1.5 | 14.4 ± 1.6 | 71.5% ± 5.4% | 78.8% ± 4.8% |
| **Hyperglycemic (>180 mg/dl)** | 36 (11.6%) | 14.0 ± 3.0 | 29.4 ± 7.6 | 64.9% ± 15.6% | 81.1% ± 12.8% |

### 3.4 Clarke Error Grid analysis

We also performed a Clarke Error Grid analysis of our results. Developed in 1987 by Clarke et al.^14^, a Clarke Error Grid is a graphical representation used to assess the clinical accuracy of blood glucose measurement systems. It is a two-dimensional representation of the accuracy of the predictions, in which the x-axis represents the reference values and the y-axis represents the values measured by the blood glucose meter being evaluated (in this case, the Bio-RFID predictions). The lines on the grid demarcate five BG zones (A to E).

The Clarke Error Grid analysis resulted in 246 of 311 (79.1%) of the blood glucose values falling into Zone A, 63 of 311 (20.25%) of the values in Zone B, 0% in Zone C, 2 of 311 (0.6%) of the values in Zone D, and 0 in Zone E, as shown in Figure 3(a).

**Figure 3(a):**
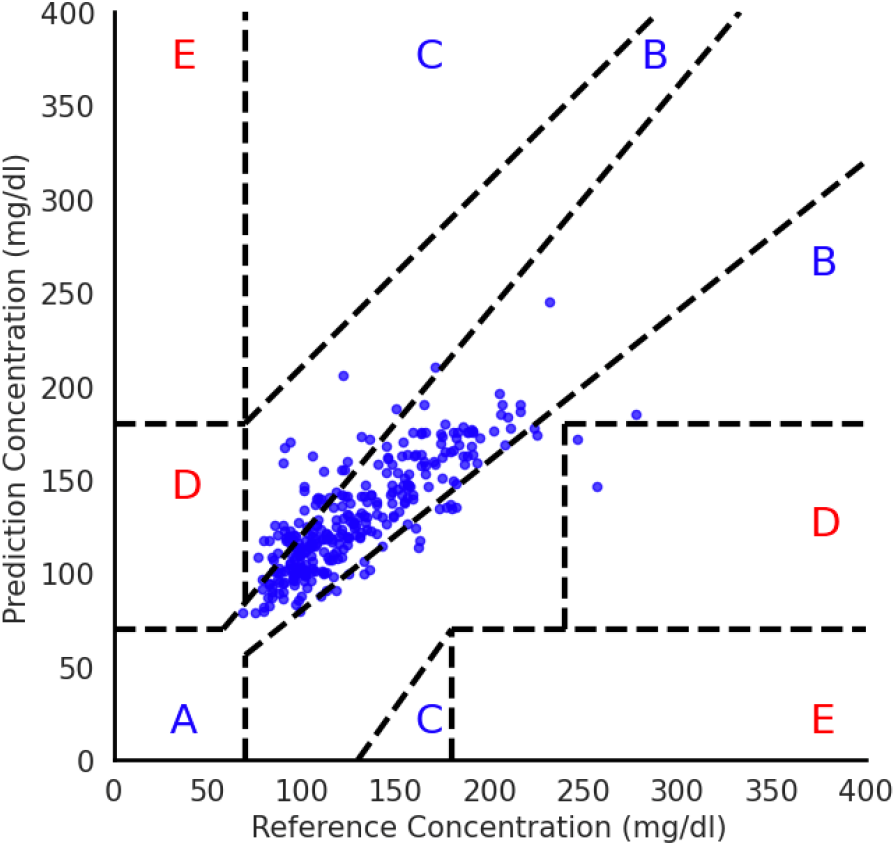
The Clarke Error Grid Analysis is depicted here to demonstrate which values in the test set fell into each error zone based on the glycemic range.

### 3.5 Surveillance Error Grid Analysis

A more modern error analysis grid that uses a finer resolution to assess the potential clinical impact of errors made by a predictive glucose model is the Surveillance Error Grid (SEG). The SEG draws on the experience of 206 diabetes clinicians to assess the potential risk of varying error. The SEG analysis of our results is shown in Figure 3(b).

**Figure 3(b):**
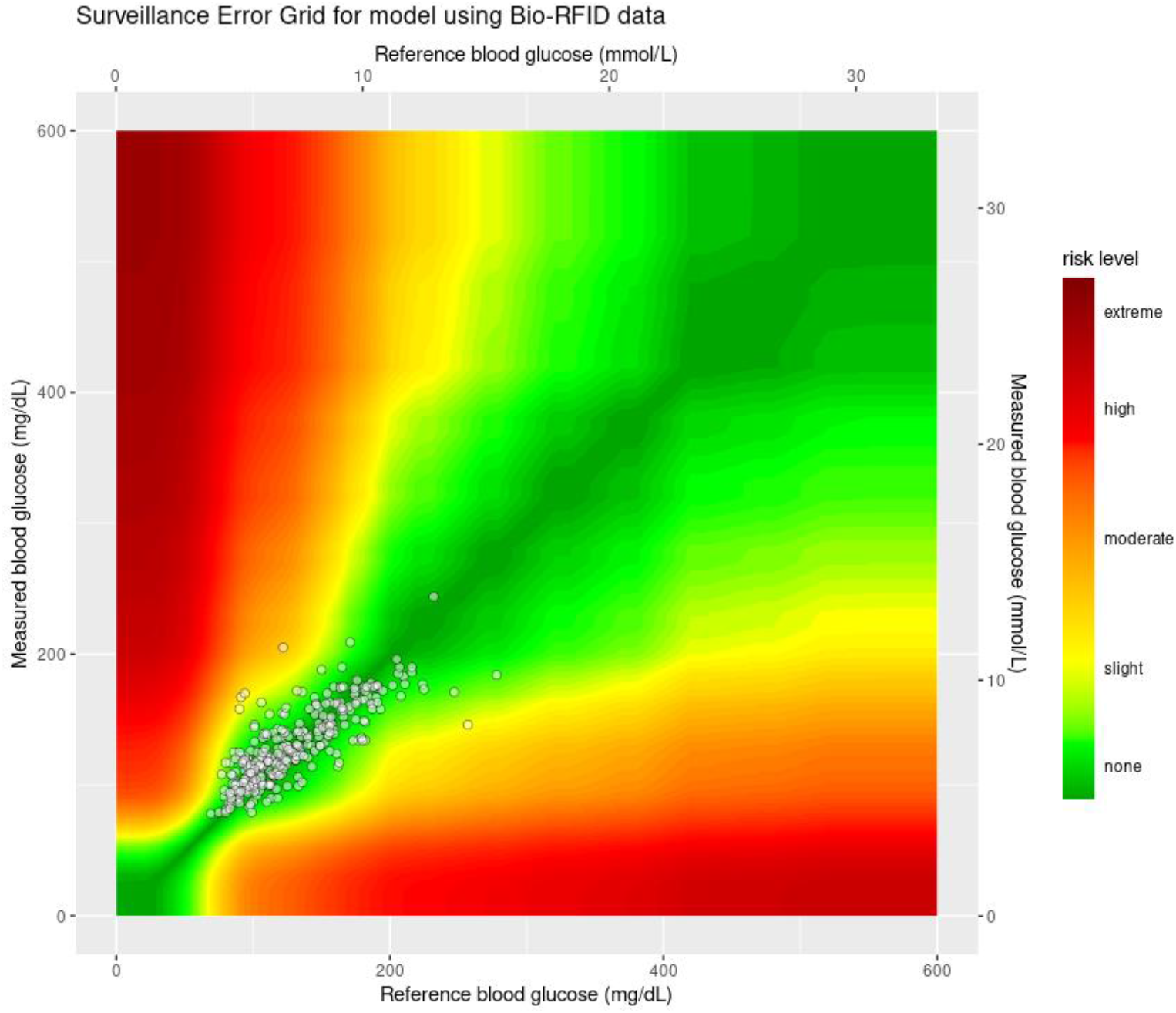
A Surveillance Error Grid analysis of model accuracy

## 4. Discussion

This work demonstrates a test in which the patented Bio-RFID sensor was able to predict blood glucose continuously and non-invasively with a MARD of 12.9%, using the output of a Dexcom G6 CGM as a reference for the measurement of blood glucose. We describe the development of a method that combines novel aspects of data collection utilizing the Bio-RFID sensor and Machine Learning techniques in the form of a lightGBM model. Data were continually collected over a 2 – 3 hour period, sampling thousands of frequencies during every 22-second sweep. In total, the sensor collected as many as 4.9 million datapoints during each test. Varying dielectric responses of glucose at different frequencies allowed the model to combine information from these frequencies to make accurate predictions.

In the test dataset, containing 311 observations, the model predicted BG with a MARD of 12.9%, which is similar to the accuracy of commercially available CGMs (see Tables 4 and 5). This was significantly higher than empirical chance on the same test set, which had a MARD of 29.8%. The model performed best in the normoglycemic range (between 70-180 mg/dL), which is not surprising given that the large majority (88.1%) of the training and test datasets contained normoglycemic values. Blood glucose estimations in the hyperglycemic range (>180 mg/dL) were still better than chance, but were worse than the normoglycemic range. We were not able to assess values in the hypoglycemic range in this study due to the lack of data in this range, likely due to the fact that all participants were healthy adults. Overall, these promising results suggest the Bio-RFID sensor can consistently and accurately measure a user’s blood glucose non-invasively.

**Table 4:** Independently validated MARD values for commercially available CGM Devices.

| Device | MARD |
| --- | --- |
| Dexcom G6 | 12.8%<sup>17</sup> |
| Senseonics Eversense | 14.8%<sup>18</sup> |
| Dexcom G5 | 16.3%<sup>18</sup> |
| Abbott Libre Pro | 18.0%<sup>18</sup> |

**Table 5:** Reported MARD values for FDA-approved CGM Devices (data from^19^).

| Device | Year of FDA Approval | MARD |
| --- | --- | --- |
| MiniMed 508 | 1990 | 25.0% |
| Paradigm | 2003 | 17.0% |
| Guardian | 2005 | 17.0% |
| Seven | 2007 | 17.0% |
| Paradigm | 2008 | 12.8% |
| Paradigm Revel | 2010 | 16.6% |
| SevenPlus | 2010 | 16.0% |
| G4 Platinum | 2012 | 13.0% |
| Enlite | 2013 | 13.6% |
| G4P 505 | 2014 | 10.0% |
| Libre | 2015 | 11.4% |
| 670G | 2016 | 9.6% |
| G5 Mobile | 2016 | 9.0% |
| Libre 14 | 2018 | 10.1% |
| G6 | 2018 | 9.0% |
| Libre 2 | 2020 | 9.2% |

A particular feature of this work that is worth emphasizing is the novelty of the sensor used in collecting data. Typically, antennas in most communication systems are designed to radiate efficiently into free space, and as such, are typically designed to be resonant structures with a specific frequency of operation and radiation pattern in mind. The Bio-RFID antenna used in this study, however, is not designed to radiate signals; instead, it is an array of elements loosely coupled to each other, where each element is located primarily in the near field of another element in the array. The loosely coupled aspect creates reasonably spaced elements, which ensures that the fields between them occupy the space in “front” of the array. This spacing is critical as it impacts coupling between the elements. Additionally, because the system operates over a broad frequency range, designing for a specific resonant frequency is not necessary, and designing for efficient coupling of fields into free space for the purposes of propagation is actually contrary to the system’s goals of coupling energy from one element to another through a material medium.

### 4.1 Comparison to independently validated clinical trials of CGMs

It is interesting to compare our MARD of 12.9% to the MARD of commercially available CGMs. Table 4 gives MARDs of some of these, as reported by published studies performed independent of the manufacturer.

In the context of these values, a MARD of 12.9% at this stage of development is quite encouraging.

### 4.2 Comparison to MARD values reported to the FDA by the CGM manufacturers

The MARD of CGMs has steadily improved over the last 30 years, since the first FDA-approved device (the MiniMed 508) in 1990. Note that the values in Table 5, unlike the independently validated values above, are those reported by the CGM manufacturer to the FDA for device approval.

In the context of these values, a MARD of 12.9% at this stage of development is quite encouraging.

### 4.3 Comparison to previous Know Labs results

In early 2023, a proof-of-concept study of the efficacy of the Bio-RFID sensor was performed using one participant^20^. In that study, we reported a MARD of 19.3%. We also reported preliminary analysis of the n=5 data in a technical feasibility study, where we reported a MARD of 20.6%^21^. Analysis in this study differs from the previous studies in a few ways. First, since the Dexcom G6 reference device was on the participants’ left arm, only data from the left arm was used in this analysis (data from both arms were used in the two previous studies). Since studies have shown that BG levels in the arms can be quite different^22^, we restricted this study to only the arm corresponding to the reference device. Another difference is the way the data is split into test and training datasets. In previous studies, a 60-20-20 train-test-validation split was used on the data, and data from each 3.5-hour test was put into only one of these splits. The present study treats each of the observations as independent, and withholds a random subset of 20% of these for the test dataset (subject to the stratification parameters described in Section 2.4). Given that the number of features (frequencies) gathered by our Bio-RFID sensor (10,001) was larger than the number of observations (1,555), we also employed feature reduction, averaging data from the Bio-RFID sensor in the temporal and frequency domains, which had not been done before, and which helped to reduce noise. The final way in which this work differed from previous studies is the machine learning model that was used. Rather than using a neural network model, which generally performs best when exposed to a significant amount of data, we elected to use a lightGBM model, which often performs well on smaller datasets.

### 4.4 Limitations

There are several limitations to this study. The most significant of these may be the number of participants in the study (n=5), which limited our ability to gather data from a diverse population. Because of this small sample size, we took an approach to stratify our training and test datasets by participant. While this stratification allows us to control variance in our test set to allow it to accurately represent the variance in our training set, it does limit our ability to understand to what degree these results would generalize to other participants. Additionally, because all participants were healthy and non-diabetic, we had a limited amount of data from the hyperglycemic range and insufficient data to assess the hypoglycemic range at all. Also, the data collected for each user was not equally balanced between devices, and future work could examine whether there is more information to be gained by studying the differences across devices and individuals. More analysis and data collection are required to accurately report glucose values on new users and new devices.

Our model was not designed to predict blood glucose directly, but rather to predict the values of a Dexcom G6 as a proxy for blood glucose. While our ultimate goal is to quantify blood glucose, the reference device provides an imperfect estimate of this, as an independent validation suggests its MARD is 12.8%. Moreover, there is a difference between the interstitial fluid the reference device is measuring and the more complex tissue the Bio-RFID sensor has access to, all of which suggests that part of our MARD of 12.9% may not be error, but an artifact of the real differences between what is being measured. In future work, it will be beneficial to explore the capability of the sensor to measure an entire cross-section of tissue, which could include arterial glucose, venous glucose, capillary glucose, interstitial glucose, and intracellular glucose. Moreover, further studies could eventually compare the Bio-RFID against a more precise blood glucose reference device, such as a YSI.

## 5. Conclusion

Overall, these results suggest that blood glucose values can be accurately estimated from the Know Labs RF-based device. Further investigations are merited to assess whether the performance can be extended to a larger participant pool and a wider range of blood glucose values in the hypoglycemic and hyperglycemic ranges. Future clinical studies should aim to generate larger volumes of high-resolution Bio-RFID data compared with industry-leading reference device data to enable further data science and model development, and ultimately achieve the goal of developing an FDA-cleared non-invasive glucose monitoring device.

## Data Availability

Data are produced in the present study are not available due to privacy and ethical concerns.

## Author Contributions

D.K.: Conceptualization (equal); Methodology (supporting); Writing – original draft (lead); Writing – review & editing (equal)

K.C.: Investigation and Methodology (lead), Conceptualization (equal); Writing – original draft (supporting), Writing – review & editing (equal)

J.H.A.: Writing – review & editing (equal)

D.S.: Data Curation and Formal Analysis (equal), Visualization (lead)

C.W.: Data Curation and Formal Analysis (equal), Visualization (supporting) B.S.: Writing – original draft (supporting); Writing – review & editing (equal)

## Footnotes

1 A preprint of this manuscript has been posted to the medRxiv server, at: https://www.medrxiv.org/content/10.1101/2023.05.25.23290539v1. DOI: 10.1101/2023.05.25.23290539v1

